# DeepCRI: Real-time EEG-based Prognostication after Cardiac Arrest

**DOI:** 10.1101/2025.09.13.25334650

**Authors:** Michel J.A.M. van Putten, Hanneke M. Keijzer, Jeannette Hofmeijer, Pauline Buscher-Jungerhans, Nicolas Gaspard, Sarah Caroyer, Albertus Beishuizen, Marleen C. Tjepkema-Cloostermans

## Abstract

Accurate prediction of neurological outcome after cardiac arrest is essential for guiding intensive care decisions. Electroencephalography (EEG) supports prognostication; however, interpretation relies on expert judgment and is often subjective and delayed.

We developed DeepCRI, a bedside-integrated deep learning system that produces continuously updated prognostic trajectories during the first 36 hours after arrest. DeepCRI uses time-dependent decision boundaries to define good-, poor-, and gray-zone regions over time, and applies a lock-in rule that fixes classification only after sustained, concordant high-confidence evidence within a compact temporal window, thereby preventing transient threshold crossings from driving decisions.

During model development in a cohort of 522 patients, DeepCRI achieved an area under the receiver operating characteristic curve (AUC) of 0.97 at 24 h, with low calibration error (ECE=0.049). Independent validation was performed in an internal (n=219) and an external cohort (n=167). In the internal validation, DeepCRI provided lock-in classifications in 81.7% of patients, achieving 100% specificity for poor outcome with a sensitivity of 49.5%, and 95.5% sensitivity for good outcome at 73.2% specificity; 18.3% remained in the gray zone. Performance in the external validation cohort was lower: 59.9% locked, and a single false predictions reduced poor-outcome specificity to 98.4%. Post hoc analysis indicated residual EMG artifacts contributed to this false poor-outcome prediction.

By embedding DeepCRI into routine ICU EEG infrastructure, we demonstrate the technical feasibility and clinical promise of continuous, real-time AI-driven prognostication for comatose patients after cardiac arrest.

## 1 Introduction

Reliable early prediction of neurological outcome after cardiac arrest helps ICU management, including family counseling and decisions on the continuation of care. Continuous EEG (cEEG) has emerged as a valuable tool, allowing identification of patterns with high prognostic value, such as early return of physiological rhythms indicating a good outcome or burst-suppression with identical bursts indicating a poor outcome [1–8]. However, visual EEG interpretation is time-consuming, subjective, and limited by substantial interrater variability, even when standardized terminology is applied [9].

To address these limitations, several machine learning models—including deep neural networks—have been proposed to support EEG interpretation for prognostication [10–14]. While these approaches report sensitivities of 50-70% at high specificity, calibration is often neglected, and predictions are typically restricted to a single, fixed time point — usually 24 h post-arrest. Such static predictions fail to capture the dynamic evolution of post-anoxic encephalopathy in individual patients. Moreover, none of the previously proposed models have been technically implemented for bedside use, substantially limiting clinical translation.

Here, we introduce DeepCRI to overcome these barriers. DeepCRI is trained on a large, prospectively collected multicenter dataset, incorporates explicit calibration, and generates continuously updated prognostic estimates over the first 36 h after cardiac arrest. A dynamic lock-in mechanism ensures that once predictions become sufficiently stable, they are fixed and no longer subject to fluctuations. This enables individualized timing of prognostication and prevents oscillatory classification. In this study, we describe the development of DeepCRI, evaluate its performance in independent validation cohorts, and demonstrate its technical integration into routine ICU EEG monitoring.

## 2 Methods

### 2.1 Study design and participants

This retrospective study included consecutive adult comatose patients (Glasgow Coma Scale ≤ 8) after cardiac arrest, admitted to the ICUs of two teaching hospitals in the Netherlands (Medisch Spectrum Twente; Rijnstate Hospital) and Erasmus Hospital (Brussels, Belgium). Sedation followed standard ICU protocols, typically propofol and, when needed, remifentanil. Clinical neuroprognostication and withdrawal-of-life-sustaining-therapy decisions followed guideline-based multimodal practice and were not based on EEG alone. The Medical Ethical Committee Twente approved the protocol (K19-11) and waived informed consent because EEG monitoring and follow-up are part of standard care.

### 2.2 Training and validation cohorts

The development cohort comprised 522 cEEG recordings (2016–2022) from MST and Rijnstate. Part of these data were used in previous studies [11, 15]. Two independent validation cohorts were defined a priori: (i) an internal validation cohort of 219 patients (2023–2024) from MST and Rijnstate, and (ii) an external validation cohort of 167 patients from Erasmus Hospital, Brussels.

### 2.3 Outcome labels

Primary outcome was the Cerebral Performance Category (CPC), dichotomized as good (CPC 1–2) versus poor (CPC 3–5). In the development cohort, CPC at 3 or 6 months post-discharge was used. In the internal validation cohort, CPC at hospital discharge was used. In the external validation cohort, CPC was obtained prospectively by telephone interview. Patients without follow-up or with non-neurological causes of death (e.g., hemodynamic failure or reinfarction) were excluded.

### 2.4 EEG recording and preprocessing

EEG monitoring was initiated within 24 hours after ICU admission and continued until recovery of consciousness, death, or up to 3–5 days. EEGs were recorded using 21 scalp electrodes (10– 20 system). Signals were band-pass filtered (1–25 Hz), downsampled to 64 Hz, detrended, and segmented into non-overlapping 10-s epochs. Epochs were automatically screened for artifacts (amplitude, relative amplitude, flat channels, and high-frequency contamination). DeepCRI values were derived from epochs passing artifact screening; windows without sufficient retained epochs yielded no prediction. Additional details are provided in the Supplementary Material.

### 2.5 Model architecture

DeepCRI uses a compact convolutional neural network (CNN) for binary outcome prediction. The network comprises three convolutional blocks with ReLU activations, batch normalization, and spatial dropout, followed by a dense layer and a sigmoid output. Gaussian input noise and max pooling were used for robustness and downsampling. Further architectural details and the full hyperparameter search space are provided in the Supplementary Material.

### 2.6 Model training and hyperparameter optimization

Model development used 5-fold stratified group cross-validation on EEG recorded 24 h after cardiac arrest. Hyperparameters were optimized with Optuna [16] to maximize discrimination (ROC AUC) and improve probability calibration (expected calibration error; ECE [17]). The optimization emphasized clinically relevant operating points, in particular high-specificity rule-in for poor outcome. Training was implemented in TensorFlow/Keras with mixed precision; additional details are provided in the Supplementary Material.

### 2.7 Dynamic decision boundaries

To generate time-evolving predictions, the trained model was applied to additional time points within the first 36 h after cardiac arrest in the development cohort. For each time point, thresholds were derived to achieve 95% specificity for good outcome and 100% specificity for poor outcome. This defined three prognostic regions over time: a good-outcome region, a poor-outcome region, and an intermediate gray zone. Thresholds per hour were subsequently fitted with an exponential function, *a* + *b* exp(*−t/τ*), yielding smooth time-dependent decision boundaries.

### 2.8 Classification and lock-in

Definitive outcome classification combined the time-dependent boundaries with a dynamic lock-in rule. Lock-in required repeated, concordant DeepCRI values within a limited temporal window, preventing transient threshold crossings from driving decisions when evidence remained unstable. Once lock-in criteria were met, classification was fixed (locked-good or locked-poor). Patients not meeting lock-in criteria remained in the gray zone. Lock-in parameters were selected using the development cohort and fixed before evaluation in the validation cohorts (details in Supplementary Material).

### 2.9 Model performance and implementation

Performance was assessed using ROC AUC, sensitivity, specificity, calibration curves, and grayzone proportion. Sensitivity and specificity confidence intervals were computed using Beta distribution intervals [18]. Calibration (ECE) was evaluated for fixed-time (24 h) probabilistic outputs only, as calibration metrics do not apply to the dynamic lock-in decision process. The final network was deployed in the clinical EEG platform (NeuroCenter EEG, Clinical Science Systems, Leiden, the Netherlands), enabling real-time bedside inference in the ICU.

## 3 Results

Baseline characteristics of the training and validation cohorts are shown in Table 1.

**Table 1:** Baseline characteristics of the training cohort and the internal and external validation cohorts.

| Characteristic | Training cohort | Internal validation | External validation |
| --- | --- | --- | --- |
| Patients, n | 522 | 220 | 168 |
| Good outcome (CPC 1–2), n (%) | 240 (46.0%) | 113 (51.8%) | 62 (36.3%) |
| Poor outcome (CPC 3–5), n (%) | 282 (54.0%) | 107 (48.2%) | 106 (63.7%) |
| Age, mean ± SD (years) | 62.5 ± 13.6 | 62.8 ± 14.6 | 61.5 ± 16.0 |
| Male sex, n (%) | 398 (76.3%) | 159 (72.3%) | 111 (66.1%) |
| Female sex, n (%) | 124 (23.7%) | 61 (27.7%) | 57 (33.9%) |

During model development, DeepCRI achieved AUC=0.97 at 24 h with ECE=0.049, consistent with well-separated predicted-probability distributions (Figure 1-right panel).

**Figure 1.**
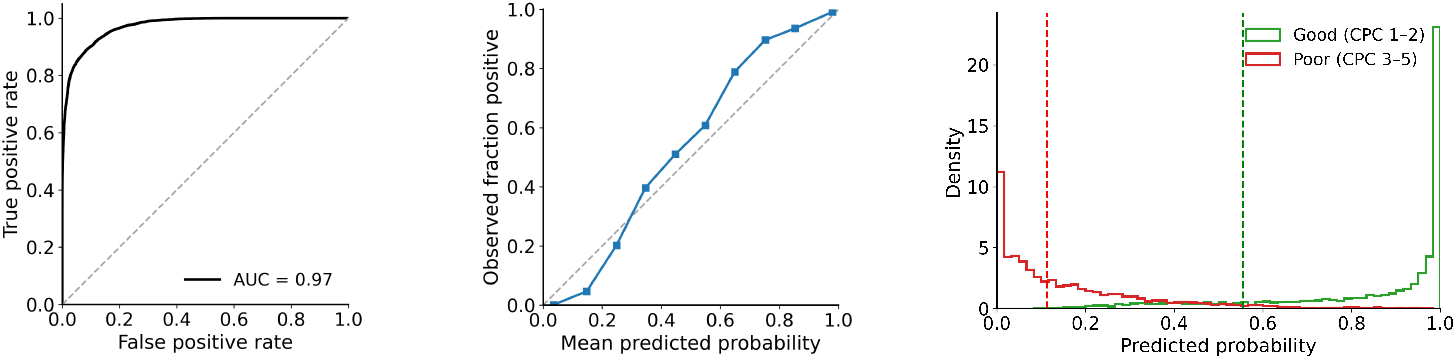
Left: ROC curve in the training cohort at t = 24 h (AUC = 0.97). Middle: calibration curve, with ECE = 0.049. Right: Predicted-probability distributions for good (green) and poor (red) outcomes. Dashed lines mark decision cut-offs chosen to achieve 100% specificity for “poor” and 95% specificity for “good”, computed on the training cohort at t = 24 h.

The time-dependent boundaries that separate the good and poor regions from the gray zone were applied unchanged in both validation cohorts (Figure 2). The parameter values of the exponential functions are in Supplementary Material.

**Figure 2.**
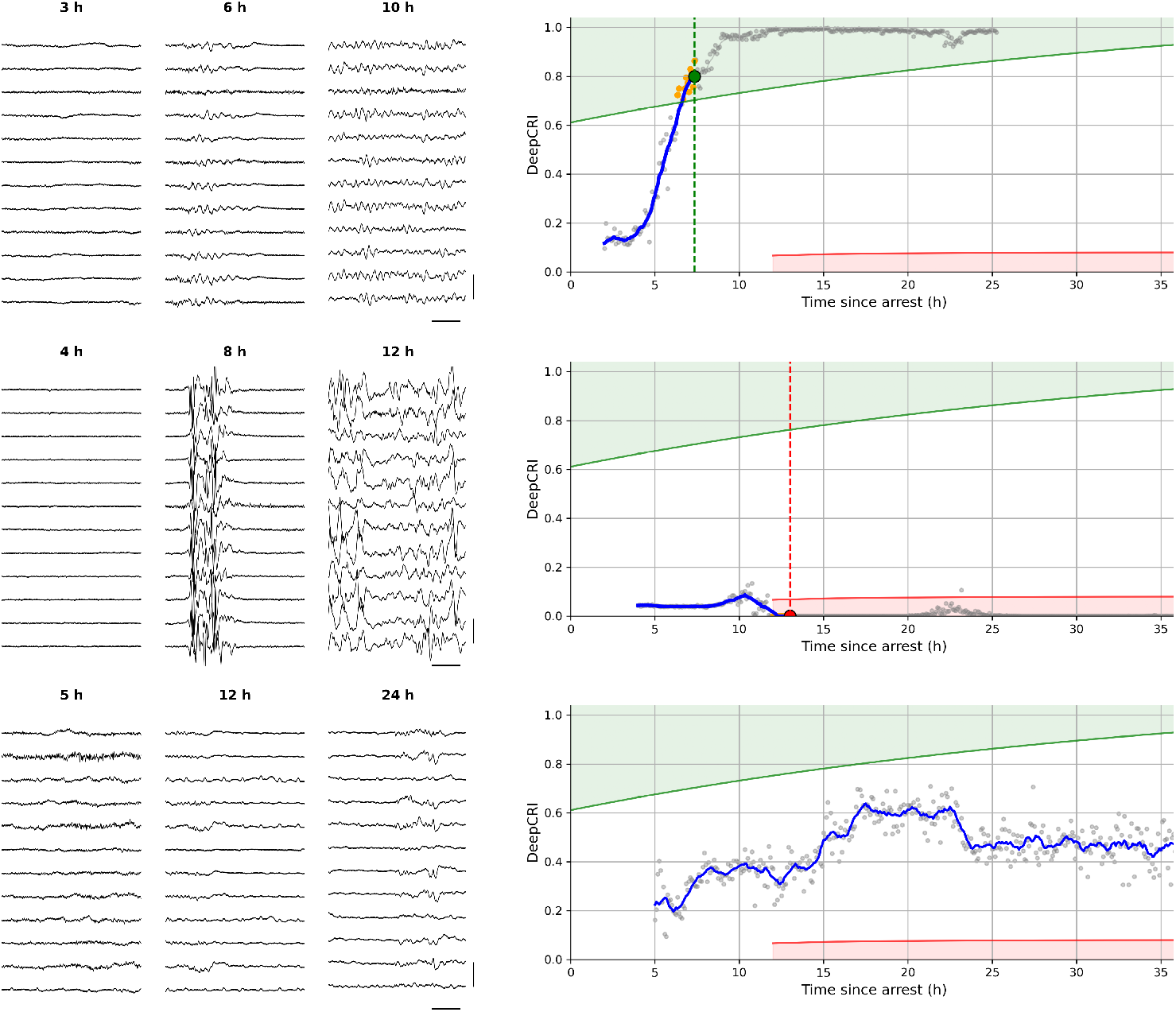
Five-second EEG epochs (left) with a reduced number of channels for display purposes and (right) corresponding DeepCRI trend curves for three patients. Times above EEG epochs indicate time since cardiac arrest. Top panel: Good outcome (CPC=1). Middle: Poor outcome (CPC=5). Bottom: DeepCRI in a patient where reliable prediction was not possible (eventually, this patient recovered well (CPC=1)). Good and poor outcome were predicted at lock-in times of 7 h and 12 h, indicated with the vertical dashed line. The dynamic boundaries for the good and poor regions are indicated with the solid green and red line, respectively. Vertical bar represents 100 µV, horizontal bar represents 1 s. EEG filter settings 1-25 Hz.

### 3.1 Lock-in parameters

Across folds, optimal parameter values showed limited variability, indicating that lock-in behavior was not dependent on fine-tuned parameter choices in the training cohort (Supplementary Table S1). Across the parameter space explored, classification behavior changed gradually, with only minor shifts in sensitivity and specificity for both poor and good outcome prediction and lock-in timing (Supplementary Table S2).

The resulting configuration required a minimum of 6 concordant DeepCRI values for good-outcome prediction and 18 values for poor-outcome prediction, evaluated within temporal aggregation windows of 140 and 90 minutes, respectively. Poor-outcome classification was additionally constrained to occur no earlier than 12.5 hours after return of spontaneous circulation (Supplementary Table S3).

### 3.2 Internal and external validation performance

Poor or good outcome classification in the internal validation cohort was achieved in 179 patients (81.7%). Sensitivity for good outcome was 95.5% (specificity 73.2%) and sensitivity for poor outcome was 49.5% (specificity 100%). Forty patients (18.3%) remained in the gray zone; these typically showed limited temporal EEG evolution. Representative trajectories illustrating good lock-in, poor lock-in, and persistent gray-zone behavior are shown in Figure 2.

In the external cohort (n=167), overall performance was lower and 100 patients (59.9%) achieved lock-in. One false poor-outcome prediction occurred. Post hoc analysis indicated abundant EMG artifacts that were not sufficiently removed by the artifact-rejection algorithm. EEG recordings also started later, and a smaller fraction of epochs survived artifact rejection (Supplementary Figure 1).

Performance on both validation sets is summarized in Table 2.

**Table 2:** Performance of DeepCRI in the internal and external validation cohorts.

| Metric | Internal validation |  | External validation |  |
| --- | --- | --- | --- | --- |
|  | Value | 95% CI | Value | 95% CI |
| Total patients | 219 | – | 167 | – |
| Patients classified (locked) | 179 (81.7%) | – | 100 (59.9%) | – |
| Gray zone patients | 40 (18.3%) | – | 64 (40.1%) | – |
| *For good outcome* |  |  |  |  |
| Sensitivity | 95.5% | 90.5–97.9% | 75.5% | 64.5–83.2% |
| Specificity | 73.2% | 61.9–82.1% | 67.8% | 55.0–78.3% |
| *For poor outcome* |  |  |  |  |
| Sensitivity | 49.5% | 40.1–59.0% | 37.4% | 28.8–46.9% |
| Specificity | 100.0% | 97.1–100.0% | 98.4% | 91.3–99.6% |
| False predictions of good outcome | 19 | – | 19 | – |
| False predictions of poor outcome | 0 | – | 1 | – |

In each cohort, one patient was excluded by the artifact detection algorithm. Without automated artifact screening, optimal lock-in parameters shifted modestly, with a slight increase in false predictions of good outcome and a increase in false predictions of poor outcome in the external cohort (Supplementary Table S4).

### 3.3 Lock-in times and bedside integration

Median lock-in times ranged from 13-18 h (good outcome) and 15-20 h (poor outcome) across cohorts (Figure 3). DeepCRI has been integrated into the clinical EEG software (NeuroCenter EEG) and is currently being evaluated in the ICU (Supplementary video).

**Figure 3.**
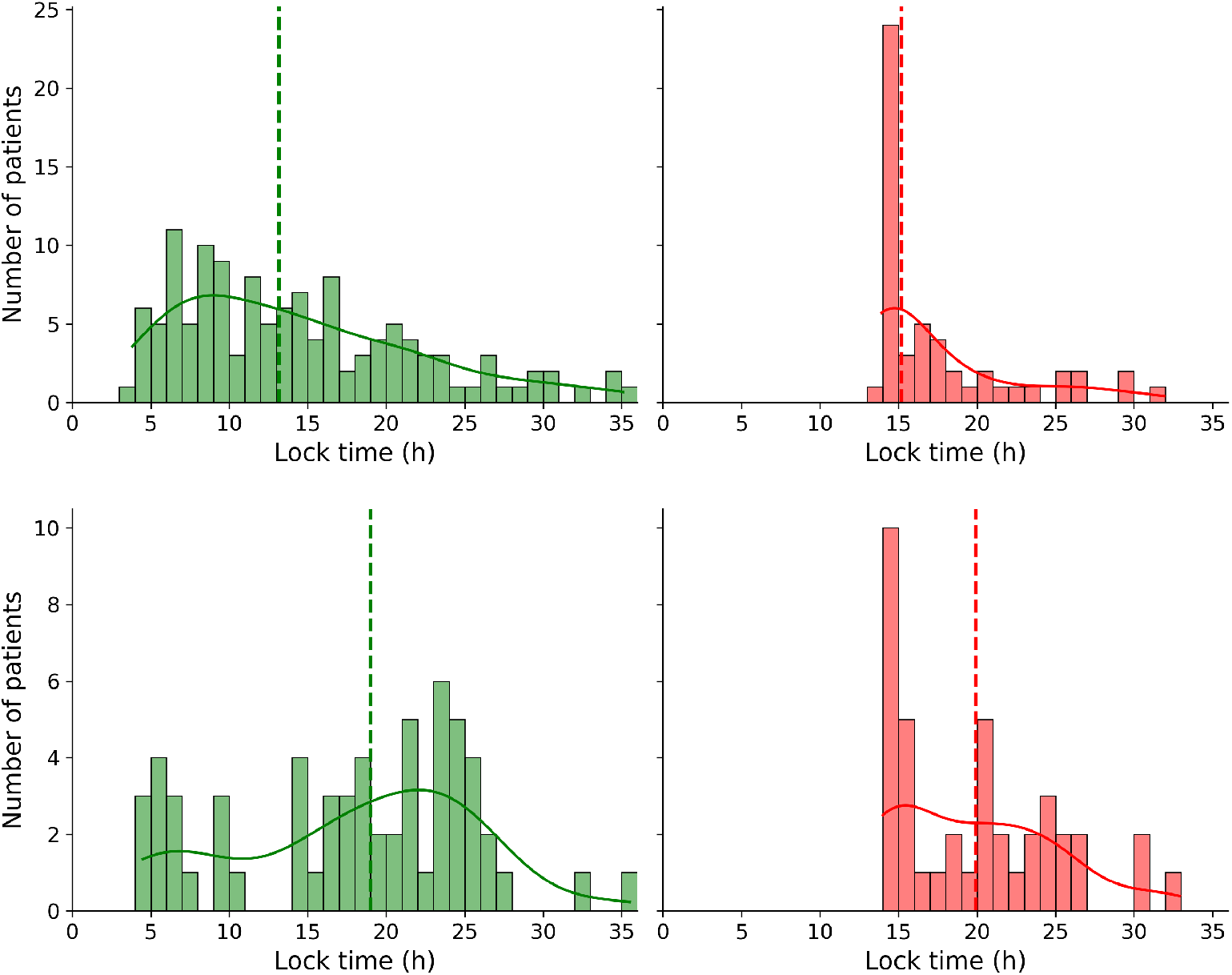
Histograms of lock-in times for patients with a good (left) and poor (right) outcome. Top: internal validation cohort. Bottom: external validation cohort. The smooth curves represent a kernel density estimate (KDE), highlighting the distribution shape and its mode. Dashed lines indicate the median lock-in times. In the internal validation, median lock-in times were 13 h for good and 15 h for poor outcome. In the external validation, these were 18 h and 20 h for good and poor outcome, respectively.

## 4 Discussion

DeepCRI is a calibrated deep learning model for EEG-based prognostication of neurological outcome in comatose patients after cardiac arrest, integrated directly into continuous bedside EEG. Unlike traditional approaches that provide static predictions, DeepCRI delivers dynamic predictions by tracking a continuously updated trajectory of EEG dynamics over time, enabling individualized timing of prognostication.

In the validation cohorts, DeepCRI yielded classifications for 60-82% of patients, achieving performance that equals or exceeds prior approaches [7, 12, 14, 19, 20], reporting sensitivities of 50–60% for poor outcome (at 95-99% specificity) and 60–75% for good outcome (at 95% specificity). External validation showed modestly reduced performance, including one false poor-outcome prediction attributable to residual EMG contamination. Our dedicated artifact-rejection algorithm based on traditional feature extraction methods may benefit from improvement. In future iterations, deep learning approaches, such as transformers or mixture-of-experts frameworks, could be explored, building on recent advances that demonstrate enhanced performance in artifact removal [21, 22]. The higher gray-zone percentage in the external validation cohort likely reflected later EEG initiation and a higher proportion of windows without sufficient epochs passing artifact-screening.

The lock-in mechanism is biologically motivated by the pathophysiology of postanoxic encephalopathy: transitions from a persistently poor EEG pattern to a sustained good pattern, or vice versa, are rare once a stable prognostic state has been reached. By requiring repeated, concordant high-confidence predictions within a limited temporal window, the lock-in mechanism aligns classification with this biological irreversibility. It reduces the risk of premature decisions driven by transient EEG fluctuations.

Median lock-in times were all within 24 hours, confirming prior evidence that early EEG contains strongly prognostic information [1, 23] and that sedation does not substantially reduce its predictive value [24]. Recent work provides additional converging evidence that most EEG-based prognostic information is captured within the first 24 hours after cardiac arrest [25]. Early EEG (within 24 hours after cardiac arrest) apparently reflects the extent of irreversible neuronal injury more clearly than later recordings, when network reorganization can obscure the underlying pathology. For instance, highly pathological patterns, such as burst suppression with identical bursts, often evolve into periodic discharges that may eventually transition to a continuous background EEG [5, 26], without clinical improvement, thereby reducing prognostic value over time.

DeepCRI is intended as an adjunct to multimodal prognostication, aligning with international guidelines [27]. DeepCRI can facilitate bedside EEG interpretation for intensivists by summarizing complex dynamics into an interpretable trajectory. This is particularly valuable in hospitals where specialized neurophysiology resources may be limited, thereby facilitating broader access to advanced prognostic information.

In future work, DeepCRI could be extended to integrate EEG with clinical examination, imaging, and biomarkers such as neuron-specific enolase (NSE) and neurofilament light (NfL) [28–31], potentially reducing uncertainty in family counseling and supporting ICU resource allocation. Its early, high-confidence predictions may enable tailored clinical decisions, such as adjusting sedation protocols for patients with favorable EEG patterns. The ongoing SELECT trial (NCT06048796) is evaluating whether early EEG-informed individualized intensive care interventions can improve post-arrest outcomes.

Our study has limitations. As in all prognostication studies, the risk of self-fulfilling prophecy must be considered. In our cohorts, however, EEG was never used in isolation to guide treatment withdrawal. Since 2019, Dutch and Belgian guidelines recommend early EEG assessment after cardiac arrest, but always in combination with clinical examination and other modalities [32]. A poor outcome determination requires, among other factors, a persistently low Glasgow Coma Scale (GCS *<* 7) after complete cessation of sedatives, typically assessed at 36 hours or later. A second limitation is that, in the internal validation cohort, the outcome was defined retrospectively by CPC at hospital discharge rather than prospectively at 3–6 months. However, discharge CPC has been shown to correlate strongly with long-term outcome [33]. Finally, model explainability remains limited, though clinicians are presented with both the real-time EEG and the DeepCRI trend curve, enabling expert verification of the raw data and artifacts.

In conclusion, DeepCRI provides robust, real-time predictions of neurological outcomes in comatose patients within 12–24 h after arrest, while explicitly allowing refraining from prognostication when reliable classification is not possible. Its dynamic confidence trajectories and lock-in mechanism reflect the evolving nature of post-anoxic encephalopathy and allow individualized timing of prognostication. DeepCRI is intended to support—not replace—expert EEG interpretation and guideline-recommended multimodal prognostication. In the external validation cohort, the false-positive poor-outcome prediction was attributable to residual EMG contamination, underscoring the importance of robust artifact handling in real-world EEG recordings. Future work will focus on improving EMG detection and mitigation strategies. By integrating DeepCRI directly into bedside EEG software, we demonstrate the technical feasibility and clinical promise of AI-driven prognostication in intensive care medicine. Ongoing prospective studies will further assess clinical impact, usability, and ethical implications.

## Data Availability

Data can be made available upon reasonable request from the corresponding author.

## Author contributions

MvP: conceptualisation, coding, data analysis, writing first draft, and data collection; MT: conceptualization, coding, data analysis, and data collection; HK: data collection; NG: data collection; AB: data collection; JH: data collection; PB: data collection; All authors: critical reading and commenting on the first draft. All authors reviewed the manuscript.

## Acknowledgements

We thank the lab technicians of the participating centers for their assistance in recording the EEGs. Clinical Science Systems is acknowledged for creating the videos.

## Ethics statement

This study was approved by the Medical Ethical Committee Twente (approval number K19-11), and all ethical guidelines were followed.

