## Supplementary Material for "DeepCRI: Real-time EEG-based Prognostication after Cardiac Arrest"

### S1 EEG preprocessing and Training

Raw EEGs were recorded using the international 10–20 system at sampling rates of 500 or 512 Hz (MST and Rijnstate) and 250 or 256 Hz (Brussels). Signals were filtered with a 1–25 Hz FIR bandpass filter and downsampled to 64 Hz. Data were detrended and segmented into non-overlapping 10-second epochs. To ensure invariance to potential EEG polarity inversions (180° phase shifts), each epoch was analyzed in both its original and sign-reversed form, and predictions were averaged over the two versions.

All EEG epochs were automatically screened for artifacts using four complementary detectors: absolute amplitude (AMPabs), relative amplitude across channels (AMPrel), flat channels (FLT), and high-frequency contamination (FRQ; EMG/alpha ratio). Each 10-s segment was flagged if any channel exceeded predefined thresholds (AMPabs $> 400$ µV, AMPrel $> 3$, FLT $> 0.8$, FRQ $> 0.3$). Only windows with at least 15 segments (out of 30) that passed the artifact-screening contributed to a DeepCRI value. If insufficient clean data were available, no DeepCRI was computed for that time window.

EEG data quality was quantified post hoc as the mean number of valid EEG segments retained after automated artifact rejection, with artifact rejection and quantification restricted to EEGs in which recording began within the first 24 h after cardiac arrest.

#### S1.1 Model architecture

The CNN comprised 3 convolutional layers with ReLU activations, batch normalization after each, and spatial dropout (fixed at 0.2). This was followed by flattening, one dense layer (ReLU), dropout (0.3–0.7), and a sigmoid output layer for binary classification. Gaussian noise (0.05) was added to the input for robustness, with zero-padding before the first convolution and max pooling after the second and third convolutions. Kernel sizes were fixed at (3×3). Hyperparameters tuned via Optuna included convolutional filters (32–128 step 32 for layers 1 and 2; 64–256 step 64 for layer 3), dense units (64–512 step 64), L2 regularization ($10^{-6}$ to $10^{-2}$, log scale), and learning rates ($10^{-6}$ to $10^{-3}$, log scale). The Adam optimizer was used with early stopping (patience=10) and learning rate reduction on plateau (factor=0.2, patience=5). A custom loss function combined binary cross-entropy with tunable penalties for calibration error (weight 0.05–0.5 step 0.05) and separation of probability distributions (weight 0.1–1.0 step 0.1), plus a fixed specificity penalty (weight 1.0). The final model was retrained using the best hyperparameters found by Optuna. For deployment, the network was recompiled with standard binary cross-entropy loss to allow inference without custom loss definitions.

#### S1.2 Model training and optimization

Model training employed 5-fold stratified group cross-validation on EEG data recorded 24 hours after cardiac arrest. Hyperparameters were optimized with Optuna [1] to maximize the area under the ROC curve (AUC) while minimizing calibration error (ECE), as described in [2], using a custom loss function incorporating specificity and separation penalties. The custom loss function combined binary cross-entropy for classification with an ECE term to ensure well-calibrated probabilities, plus penalties to enforce high specificity (e.g., minimizing false positives for poor outcomes) and maximize separation between good/poor outcome distributions. This optimization prioritized clinically relevant thresholds, such as 100% specificity for poor predictions. The model was implemented in Python using TensorFlow/Keras with mixed precision training. GPU training was performed on an NVIDIA GTX 1080 under Rocky Linux 9. Training required approximately 5 days.

### S2 Dynamic boundaries

For the boundary of the good-outcome region, we found

$$\mathrm{Thr}_{\mathrm{good}}(t) = 1.13–0.52\exp(–t/37.8) \tag{1}$$

and for the poor-outcome

$$\mathrm{Thr}_{\mathrm{poor}}(t) = 0.08–0.10\exp(–t/5.7) \tag{2}$$

where t is time in hours. Values were clipped to [0, 1].

### S3 Lock-in parameters

A data-driven Optuna search was used to tune five lock-in parameters: (i) the minimum number of concordant DeepCRI values, (ii) their temporal compactness within an aggregation window, and (iii) outcome-specific timing constraints (including the earliest allowed poor-outcome lock-in after return of spontaneous circulation). The objective maximized sensitivity under constraint of 100% specificity for poor-outcome prediction. Parameter ranges are listed in Table S1.

Table S1: Sensitivity analysis of lock-in parameters based on Optuna optimization trials using the training cohort.

| **Parameter** | **Explored range** | **Median** | **Near-optimal range** |
|---|---|---|---|
| min_good | 6–20 | 19 | 14–20 |
| min_poor | 6–20 | 16 | 13–18 |
| good_win (min) | 20–180 | 131 | 105–153 |
| poor_win (min) | 20–180 | 144 | 74–163 |
| poor_hour (h) | 6–18.0 | 12.6 | 10.8–13.5 |

Across cross-validation folds within the training cohort, sensitivity and specificity varied only modestly, confirming that classification performance was robust to variation in lock-in parameter selection (Table S2).

Table S2: Cross-validated classification performance across five folds within the training cohort (n = 522). Values are reported as mean ± standard deviation, illustrating robustness of classification performance to fold-wise variation in lock-in parameter selection.

| **Metric** | **Mean ± SD** |
|---|---|
| Sensitivity (good outcome) | 84.5 ± 2.0% |
| Specificity (good outcome) | 81.4 ± 9.2% |
| Sensitivity (poor outcome) | 46.3 ± 7.1% |
| Specificity (poor outcome) | 99.0 ± 1.4% |
| Gray-zone fraction | 0.230 ± 0.030 |

Median values in Table S1 may differ slightly as the final lock-in parameters were obtained using all data in the training cohort (Table S3).

Table S3: Final lock-in parameters using the full training cohort.

| Parameter | Final value |
|---|---|
| Minimum DeepCRI values (good outcome) | 6 |
| Minimum DeepCRI values (poor outcome) | 18 |
| Temporal window, good outcome | 140 min |
| Temporal window, poor outcome | 90 min |
| Minimum time for poor-outcome lock-in | 12.5 h |

These values were used to evaluate the classification performance in the validation cohorts.

### S4 Performance without artifact rejection

Performance of DeepCRI without artifact rejection is summarized in Table S4. In this analysis, the total number of patients is 220 (internal) and 168 (external), with 190 (86.4%) and 120 (71.4%) locked, respectively, and 30 (13.6%) and 48 (28.6%) remaining in the gray zone. False predictions of poor outcome remained 0 (internal) and increased to 3 (external), while false predictions of good outcome increased modestly (25 internal, 28 external). Totals differ slightly because no patients are excluded by artifact detection in this analysis.

Table S4: Performance of DeepCRI without artifact rejection (sensitivity analysis).

| | **Internal validation** | | **External validation** | |
|---|---|---|---|---|
| **Metric** | **Value** | **95% CI** | **Value** | **95% CI** |
| Total patients | 220 | – | 168 | – |
| Patients classified (locked) | 190 (86.4%) | – | 120 (71.4%) | – |
| Gray zone patients | 30 (13.6%) | – | 48 (28.6%) | – |
| *For good outcome* | | | | |
| Sensitivity | 97.1% | 92.9–98.8% | 78.7% | 69.9–85.9% |
| Specificity | 68.8% | 57.9–77.9% | 64.1% | 53.0–73.9% |
| *For poor outcome* | | | | |
| Sensitivity | 51.9% | 42.5–61.2% | 45.5% | 36.5–54.8% |
| Specificity | 100.0% | 97.3–100.0% | 95.9% | 88.6–98.5% |
| False predictions of good outcome | 25 | – | 28 | – |
| False predictions of poor outcome | 0 | – | 3 | – |

### S5 Start of EEG and artifacts

The distribution of EEG initiation times and EEG data quality after automated artifact rejection across the three participating centers is shown in Figure S1.

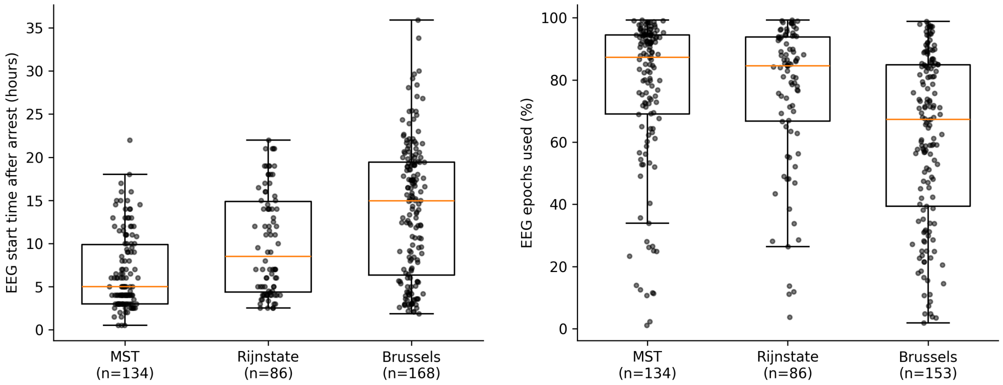


Figure S1: Left: Distribution of cEEG initiation times relative to cardiac arrest across the three participating centers. Median EEG start times were 5.0 h (MST), 8.5 h (Rijnstate), and 13.3 h (Brussels). Right: Percentage of EEG data retained for DeepCRI analysis after automated artifact rejection, considering only EEG recorded within the first 24 h after cardiac arrest. Median usable EEG percentages were 87.3 (MST), 84.6% (Rijnstate), and 67.3% (Brussels).

### S6 Supplementary videos

A demonstration of DeepCRI integrated into the NeuroCenter EEG platform is provided as Supplementary video S1.
